# Geographic and demographic heterogeneity of SARS-CoV-2 diagnostic testing in Illinois, USA, March to December 2020

**DOI:** 10.1101/2021.04.14.21255476

**Authors:** Tobias M Holden, Reese A.K. Richardson, Philip Arevalo, Wayne A. Duffus, Manuela Runge, Elena Whitney, Leslie Wise, Ngozi O. Ezike, Sarah Patrick, Sarah Cobey, Jaline Gerardin

**Affiliations:** Northwestern University Feinberg School of Medicine, Chicago IL; Department of Chemical and Biological Engineering, Northwestern University, Evanston IL; Department of Ecology and Evolutionary Biology, University of Chicago, Chicago IL; Center for Preparedness and Response, Division of State and Local Readiness, Centers for Disease Control and Prevention, Atlanta GA; Illinois Department of Public Health, Springfield IL; Department of Preventive Medicine and Institute for Global Health, Northwestern University, Chicago IL

**Keywords:** SARS-CoV-2, COVID-19, diagnostic testing, racial disparities, case fatality rate, infection fatality rate, Illinois

## Abstract

**Background:** Availability of SARS-CoV-2 testing in the United States (U.S.) has fluctuated through the course of the COVID-19 pandemic, including in the U.S. state of Illinois. Despite substantial ramp-up in test volume, access to SARS-CoV-2 testing remains limited, heterogeneous, and insufficient to control spread.

**Methods:** We compared SARS-CoV-2 testing rates across geographic regions, over time, and by demographic characteristics (i.e., age and racial/ethnic groups) in Illinois during March through December 2020. We compared age-matched case fatality ratios and infection fatality ratios through time to estimate the fraction of SARS-CoV-2 infections that have been detected through diagnostic testing.

**Results:** By the end of 2020, initial geographic differences in testing rates had closed substantially. Case fatality ratios were higher in non-Hispanic Black and Hispanic/Latino populations in Illinois relative to non-Hispanic White populations, suggesting that tests were insufficient to accurately capture the true burden of COVID-19 disease in the minority populations during the initial epidemic wave. While testing disparities decreased during 2020, Hispanic/Latino populations consistently remained the least tested at 1.87 tests per 1000 population per day compared with 2.58 and 2.87 for non-Hispanic Black and non-Hispanic White populations, respectively, at the end of 2020. Despite a large expansion in testing since the beginning of the first wave of the epidemic, we estimated that over half (50-80%) of all SARS-CoV-2 infections were not detected by diagnostic testing and continued to evade surveillance.

**Conclusions:** Systematic methods for identifying relatively under-tested geographic regions and demographic groups may enable policymakers to regularly monitor and evaluate the shifting landscape of diagnostic testing, allowing officials to prioritize allocation of testing resources to reduce disparities in COVID-19 burden and eventually reduce SARS-CoV-2 transmission.

## Background

As of December 2020, more than 95 million cases of SARS-CoV-2 infection had been detected globally in more than 190 different countries and territories (1). Yet, those 95 million cases were estimated to be a small fraction of all SARS-CoV-2 infections, with the true number of infections likely to be at least an order of magnitude higher (2). The United States (U.S.) has been hit hard by COVID-19, and limited access to diagnostic tests early in the pandemic likely contributed to substantial community spread prior to the implementation of stay-at-home policies (3). While testing in the U.S. expanded enormously after March 2020, access to testing remained uneven: per capita testing rates varied regionally and across multiple sociodemographic factors. Within a state, some testing sites ran out of reagents by mid-week, while in other areas, employers and universities were implementing routine mass testing (4,5).

SARS-CoV-2 diagnostic testing is considered a cornerstone for containing the virus. Testing informs surveillance, which guides evidence-based decision-making on hospital resource planning, implementation and relaxation of mitigation measures, and allocation of public health resources. Testing is also a means to control virus spread. Individuals who test positive are more likely to self-isolate, reducing onward transmission (6). When testing is insufficient, surveillance quality suffers, and infectious individuals may not adequately self-isolate. Understanding fine-scale heterogeneity in testing and changes over time is essential for understanding where additional resources should be directed.

The U.S. state of Illinois, with 12.7 million residents, is the sixth most populous state and representative of the country in terms of racial demographics and income distribution (7,8). Illinois contains a major urban center in the northeast, the city of Chicago (Illinois COVID-19 Region 11), with surrounding suburban counties (Figure 1A). Another urban center is in the southwest (Region 4) adjacent to the city of St. Louis in the neighboring state of Missouri. The remainder of the state is primarily rural. Using aggregate testing data from the Illinois Department of Public Health (IDPH), census data, and individual-level case and death data from IDPH, we characterized testing rates across different regions of the state, across age groups, between racial and ethnic groups, and over time. Since infections are only identified as cases upon positive diagnostic test, we assessed whether case fatality ratios (CFR) might serve as a crude indicator for under-testing in the absence of other information and estimated the fraction of all SARS-CoV-2 infections that have been detected in Illinois.

**Figure 1.**
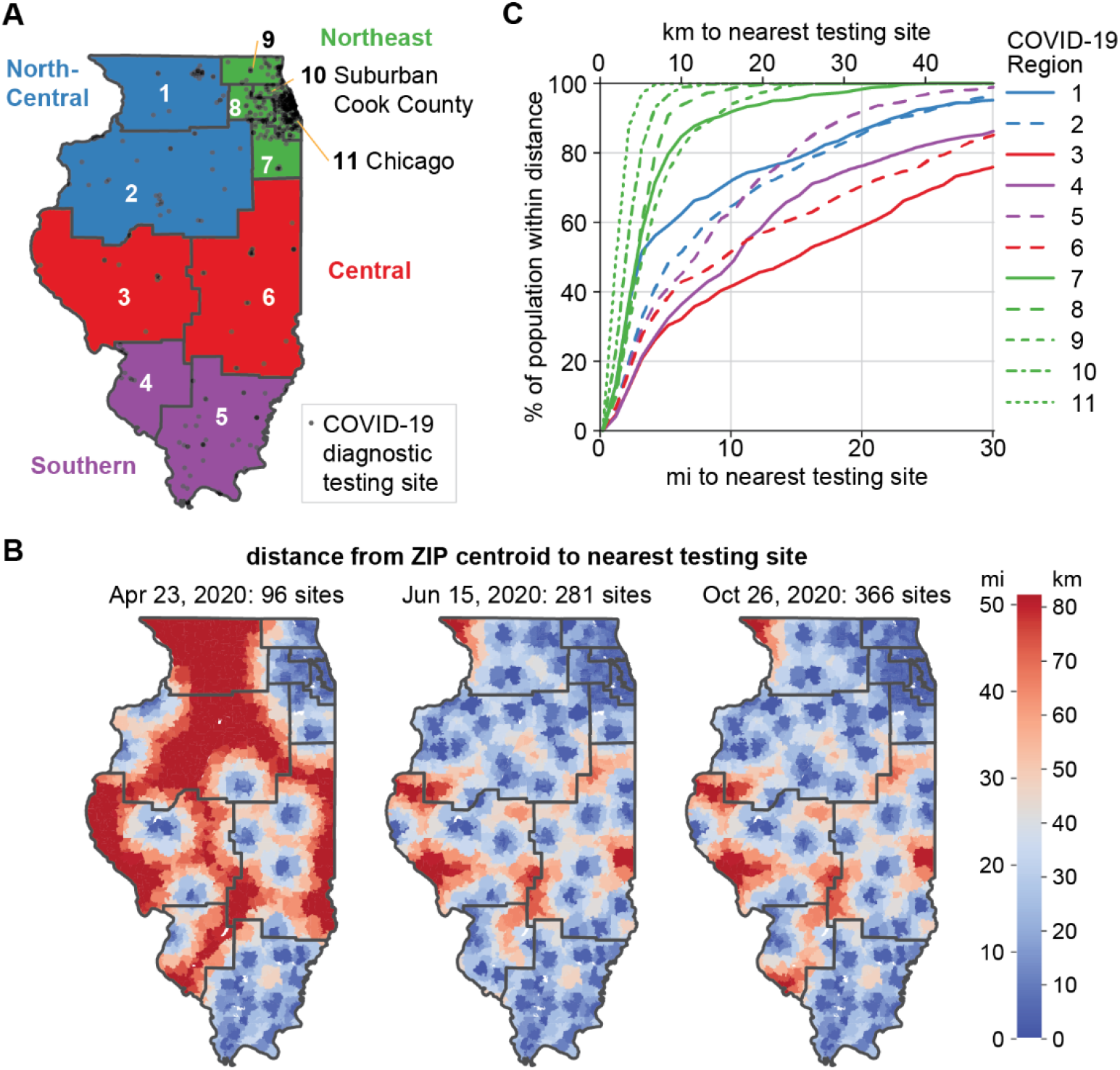
Spatial distribution of and access to COVID-19 diagnostic testing sites in Illinois. (A) State-designated COVID-19 regions (numbered) and super-regions (colored) of Illinois. Testing sites listed on IDPH’s website on October 26, 2020, are shown in transparent black. (B) Distance to nearest Illinois testing site location by ZIP code, with COVID-19 region boundaries shown in black. Distances were measured from the centroid of each ZIP code. (C) Cumulative distribution of population living within a certain distance of an Illinois testing site by COVID-19 region. Distances were measured from the centroid of each census block group to Illinois testing site locations on October 26, 2020.

## Methods

### Case definition

This work defines cases as SARS-CoV-2 infections recorded in Illinois surveillance as a result of a positive diagnostic test, regardless of symptom status.

### Datasets

County-level positive tests and total tests were obtained from the Illinois National Electronic Disease Surveillance System (I-NEDSS) database maintained by IDPH. Daily testing volume data included 12,746,960 total specimens and 1,131,284 positive specimens recorded from March 17, 2020, to December 31, 2020, stratified by age, county of test, and race/ethnicity. Moving averages of daily testing volume were calculated on a seven-day lagging window. Until October 14, 2020, only molecular tests (reverse transcriptase polymerase chain reaction [RT-PCR] tests) were reported in this dataset. On October 14, 2020, IDPH began reporting antigen tests in this dataset. Testing site locations were scraped from the IDPH website on April 23, June 15, and October 26, 2020.

Individual-level case data, including date of first positive specimen, patient’s home ZIP code, race, ethnicity, hospital admission status, and date of death, were obtained from I-NEDSS. Data were pulled on March 16, 2021, and included 1,098,549 cases reported to IDPH, 907,799 of which had specimen collection dates in 2020. Among all cases that had a date of death in I-NEDSS, 18,830 were designated as having died due to COVID-19 and were considered confirmed deaths. Cases classified as died from COVID-19 met at least one of the following criteria: presence of COVID-19 on the death certificate; death within 30 days of symptom onset/diagnosis or during hospitalization, unless the cause of death is clearly unrelated to COVID-19 (e.g. accident); never returned to baseline health after diagnosis; autopsy result consistent with COVID-19. Individuals with date of first positive specimen on or before December 31, 2020, whose deaths were confirmed in I-NEDSS after March 16, 2021, would not be included in this death tally.

In the case data, individuals were assigned to a region based on the county of symptom onset, and secondarily on listed ZIP of residence if county of onset was not available. To estimate under-reporting rates, a naïve (crude) CFR was calculated as cumulative deaths divided by cumulative cases. Counties were aggregated into COVID-19 Regions as defined by IDPH specifically for the COVID-19 response (9) and into super-regions as follows: COVID-19 Regions 1 and 2 into North Central super-region; 3 and 6 into Central super-region; 4 and 5 into Southern super-region; and 7-11 into Northeast (Figure 1A). County populations were obtained from the 2018 American Community Survey (ACS) (10).

Self-reported race or ethnicity were available for 657,219 (72.4%) individual cases reported to IDPH and were missing for the remainder. Cases with multiple races reported were categorized as “Other”. All individuals with Hispanic/Latino as ethnicity were categorized as Hispanic/Latino regardless of race(s). Individuals with “unknown” ethnicity or no reported ethnicity were considered non-Hispanic/Latino. For brevity, non-Hispanic Black and non-Hispanic White populations are referred to as Black and White, respectively.

In the testing dataset, race or ethnicity was recorded for 7,143,108 total specimens (56.0%) and 652,643 positive specimens (57.7%) using a single variable indicating either a non-Hispanic race or Hispanic-Latino ethnicity.

### Measuring distance to nearest testing site

We computed the distance from the centroid of each census block group to its nearest testing site location on October 26, 2020, and used the estimated population of each census block group (2016 ACS via Safegraph) to create a cumulative distribution of this distance over a region’s population. Census block groups were assigned to COVID-19 region by whether a census block group’s centroid fell within the boundaries of a COVID-19 region. Distance to the nearest testing site by ZIP was measured from each ZIP’s centroid to the nearest testing site location listed on IDPH’s website as sites open to the public on April 23, June 15, and October 26, 2020. The IDPH list of testing sites is not comprehensive as some testing sites asked not to be listed, and data on the actual number of sites offering testing were not available.

### Estimation of infection detection rate

To estimate the fraction of infections detected in a particular week (*f*_*inf det*_), the expected infection fatality ratio (IFR) among cases with new positive specimens collected during that week was calculated based upon these cases’ age distribution using either (i) the exponential meta-regression performed by Levin et al. (11), which used first-wave data from multiple countries; or (ii) estimates from O’Driscoll et al. (12), which uses an ensemble model incorporating data from multiple countries to infer age-specific infection mortality rates. Infection fatality ratio is the fraction of all SARS-CoV-2 infections that result in death. The results of the meta-regression of Levin et al. (11), with associated uncertainties for each coefficient, are reproduced below:

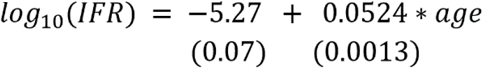

Due to the fact that, at any given time, the age distribution of cases (i.e. detected infections) was not necessarily representative of the age distribution of all incident infections, only cases 61-70 years of age were included in this analysis. This age range was selected for its large number of cases with confirmed COVID-19 deaths (>12 every week of specimen collection except for the week of March 8^th^, 2020, the first week in which a fatal case was documented in ages 61-70 years) while being less likely than older age groups (>70 years) to be associated with the less representative transmission and testing conditions in long-term care facilities. For our estimates using IFR from O’Driscoll et al. (12), IFR was uniformly sampled from 0.39 - 1.24% for ages 61-70 years, which was obtained by combining IFR of 0.46% (95% CI: 0.39 - 0.57%) for ages 60-64 years and 1.08% (95% CI: 0.92 - 1.24%) for ages 65-69 years.

First, a naive (crude) estimate of *f*_*inf det*_ was made by dividing the expected IFR by the reported CFR among that week’s cases. To account for a decreasing IFR due to improved clinical outcomes among the infected over the course of the pandemic, a second estimate was made by adjusting the expected IFR down by the relative decrease in a sigmoid curve fitted to the hospital fatality ratio (HFR) over time among people aged 61-70 years. HFR was calculated from I-NEDSS data as fraction of admitted cases that were later recorded as a death due to COVID-19. This sigmoid curve was fitted to weekly HFR with a non-linear least squares regression.

To account for unreported deaths, a third estimate was made in which the adjusted IFR was divided by the CFR, then multiplied by the estimated fraction of all COVID-19 deaths that were reported as COVID-19 deaths (*f*_*death det*_) on the median date of death among that week’s cases. To estimate *f*_*death det*_, we compared observed counts of COVID-19 deaths to excess deaths in select-cause mortality data (Figure S1). Select-cause mortality data provided by the National Center for Health Statistics (NCHS), including respiratory diseases and circulatory diseases among others, showed the expected weekly count of deaths by a selection of comorbid conditions of COVID-19 alongside the reported counts of deaths by these causes that occurred in the state in 2020 (13). Excess select-cause deaths are calculated as the difference between the expected weekly select-cause death curve and the actual weekly select-cause death curve. Assuming that all excess select-cause deaths were attributable to COVID-19 and that the epidemic did not appreciably reduce deaths indirectly due to other causes in the list of select causes curated by NCHS, we calculated *f*_*death det*_ by dividing the observed number of COVID-19 deaths each week (from I-NEDSS) by the excess select-cause deaths in the same week. To account for uncertainty in our estimate of *f*_*death det*_, we then sampled 1,000 realizations of excess deaths using a Skellam distribution, which models the difference between two Poisson random variables, and recalculated *f*_*death det*_ for each realization.

These three estimates were made from the week of March 8^th^ to the week of December 27^th^, with 1,000 bootstrapped samples taken on a weekly basis from estimates of CFR for cases in a given week, expected IFR, the prediction band of the sigmoid curve fitted to HFR, and the estimates of *f*_*death det*_ to generate a range of estimates for *f*_*inf det*_. For cases in a given week, estimates of *f*_*death det*_ were drawn from the week of the median death date of that week’s cases. All infection detection estimates, as well as HFR and *f*_*death det*_, were conducted at the statewide level.

## Results

### Spatio-temporal variation in testing and access to testing in Illinois

As of December 31, 2020, more than 900,000 SARS-CoV-2 cases and 18,000 COVID-19 deaths were recorded in Illinois (Figure 2) (14). The first wave of COVID-19 occurred in early May in the Northeast and Southern super-regions and in mid- to late-May in the Central and North-Central super-regions. COVID-19 Regions 1 and 7-11 experienced the bulk of the first-wave cases and deaths, and Regions 11 (city of Chicago) and 10 (suburban Cook County) recorded the highest peaks in daily detected cases during this time. By August 2020, daily detected cases in Regions 2, 3, 4, 5, and 6 had surpassed their peak numbers in May. By October 2020, all COVID-19 regions were experiencing a second wave of hospitalizations and deaths.

**Figure 2.**
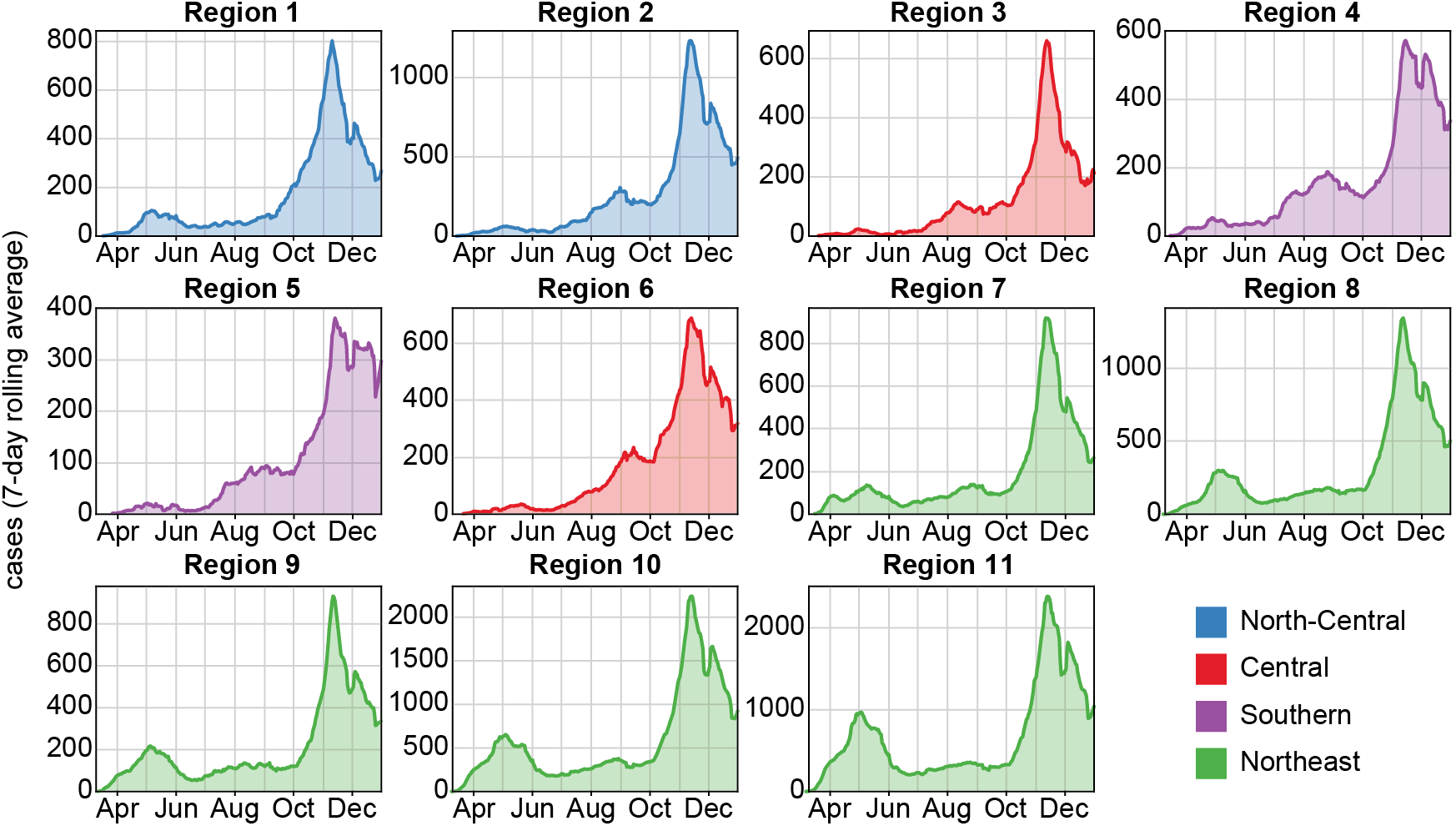
Epidemic trajectory of COVID-19 cases in the 11 COVID-19 regions of Illinois in 2020.

Reflecting both population density and the regional differences in initial burden of COVID-19, the majority of testing sites were located in the Northeast super-region (64.7% of Illinois testing sites on October 26, 2020), and many were in Regions 10 and 11 (42.1% of Illinois testing sites on October 26, 2020) (Figure 1A). Although the number of diagnostic testing sites in the state has nearly quadrupled since April (Figure 1B), most new testing sites since June have been established in the Northeast super-region. Over 50% of individuals in Regions 3, 4, and 6 resided more than 10 miles from the nearest Illinois testing site (Figure 1B and 1C). This distance is not necessarily reflective of the distance any given individual in an area must travel or will travel to receive a test. Many test sites restricted testing to symptomatic individuals, close contacts, or in-network patients in terms of referrals or insurance plans, but testing criteria data were not sufficiently available or reliable to assess access to unrestricted testing. Moreover, these restrictions were subject to continuous change as the availability of resources at individual sites fluctuated. Conversely, individuals in border areas could seek testing across state lines.

Although testing was limited in all COVID-19 regions during the first wave, testing volume expanded 5- to 10-fold between early May and the end of December (Figure 3). Controlling for population size, the overall testing rate was highest in Regions 10 and 11 during the early outbreak in March to June 2020. Some regions (particularly Regions 1 and 6) that were slower to increase testing in the first wave outpaced other regions in testing intensity by late October due to prioritization of the deployment of mobile teams to areas of greatest impact (meat processing plants, low income housing areas, etc.) and establishment of community drive through testing sites where none previously existed. In November and December there was a concerted increase in testing in all Regions, with Region 3 achieving the highest testing rates in the state, before a decrease around Christmas.

**Figure 3.**
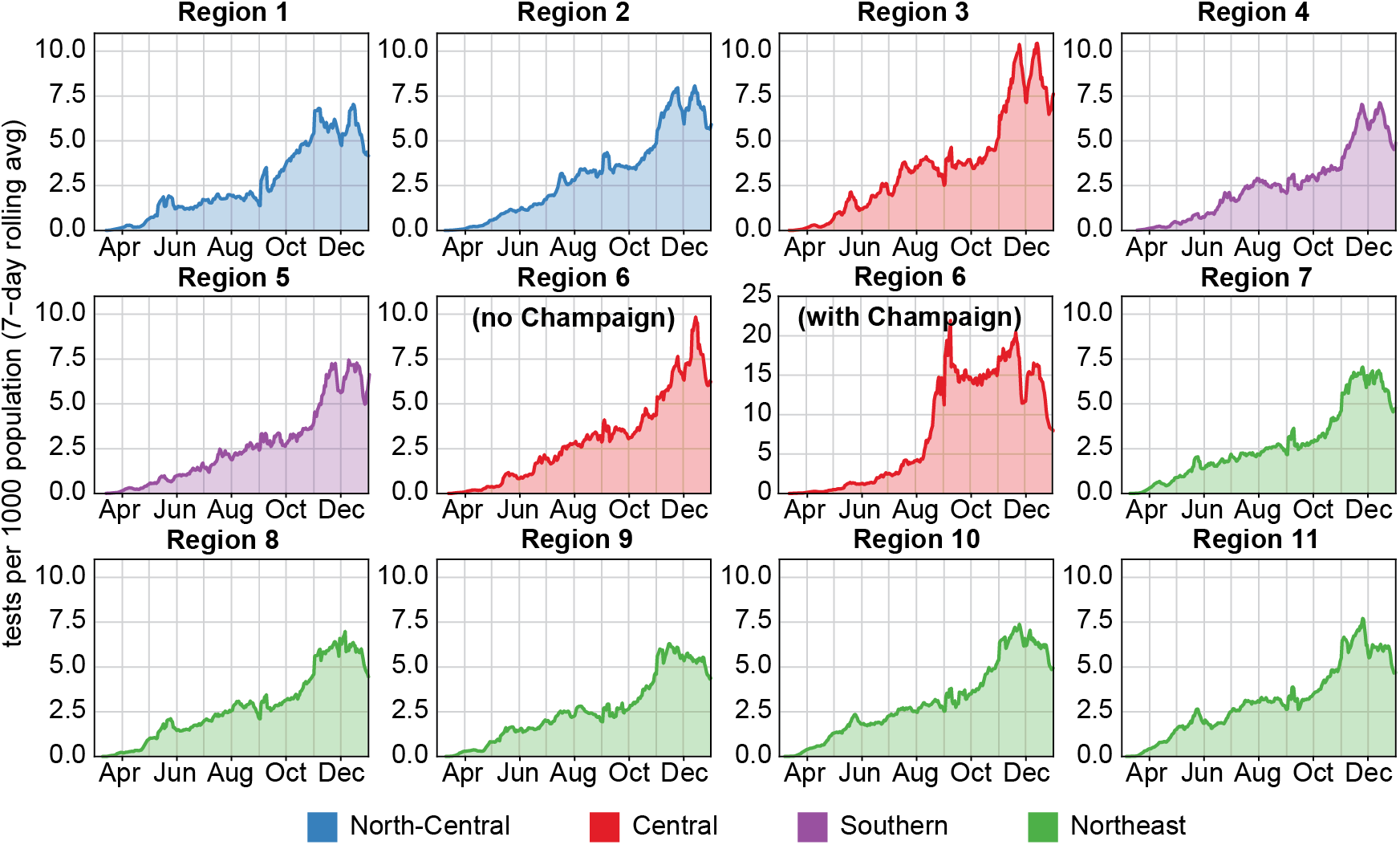
Daily SARS-CoV-2 diagnostic tests administered per 1,000 population in each COVID-19 region in Illinois in 2020. Shown: 7-day moving averages. Colors indicate super-region membership of each COVID-19 region, as indicated in Figure 1A.

In Region 6, overall testing intensity was dominated by the University of Illinois at Urbana-Champaign (UIUC) due to their efforts to conduct mass testing on their entire campus population (5) (Figure 3 Region 6 with and without Champaign County, Figure 4A). However, outside of Champaign County, the remainder of Region 6 contained some of the lowest per-capita testing in Illinois, reflecting the substantial portion of Region 6 residents who resided more than 10 miles from a testing site (Figure 1C). There was considerable county-level heterogeneity in testing intensity (Figure 4A) and positive tests per capita (Figure 4B) within Regions 1-6.

**Figure 4.**
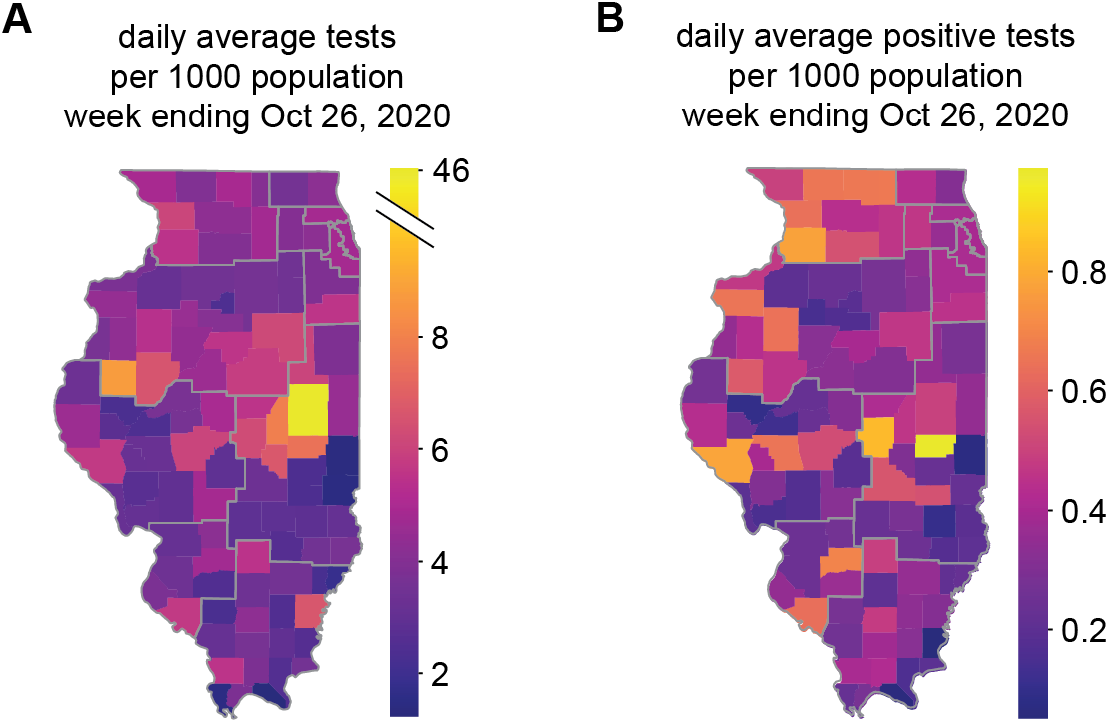
(A) County-level average daily SARS-CoV-2 diagnostic tests per 1,000 population for a representative week ending October 26, 2020. Champaign County, in COVID-19 Region 6, stands out with the highest per capita testing rate in the state. Central and Southern counties have the lowest rates. (B) County-level average daily positive tests per 1,000 population for the week ending October 26, 2020.

### Changing demographics of the tested population

Prior to mid-August, testing was most intensive in the elderly population, with those aged over 80 years receiving the most tests per capita, and intensity of testing increasing with age (Figure 5A). Routine testing in long-term care facilities contributed to higher testing intensity in the elderly population (15,16). In July, pilot testing at UIUC led testing in people aged 18-22 years to exceed testing by more than twice the rate in all other age groups, even the over 80-year-old group. Testing at other university campuses would also contribute to the increased testing rate in 18-22 year-old people, and the testing rate declined in late November following the Thanksgiving holiday and winter recess. Working-age adults may have been subject to routine testing at employers’ behest. Pediatric testing, including testing in older children, remained much lower than all other age groups. This could have been due to lower prevalence of SARS-CoV-2 infection in children because of lower susceptibility (17), low rates of test-seeking among SARS-CoV-2-infected children because they are less likely to be symptomatic, barriers to accessing testing because some providers did not test pediatric patients, or simply a lack of routine testing in children.

**Figure 5.**
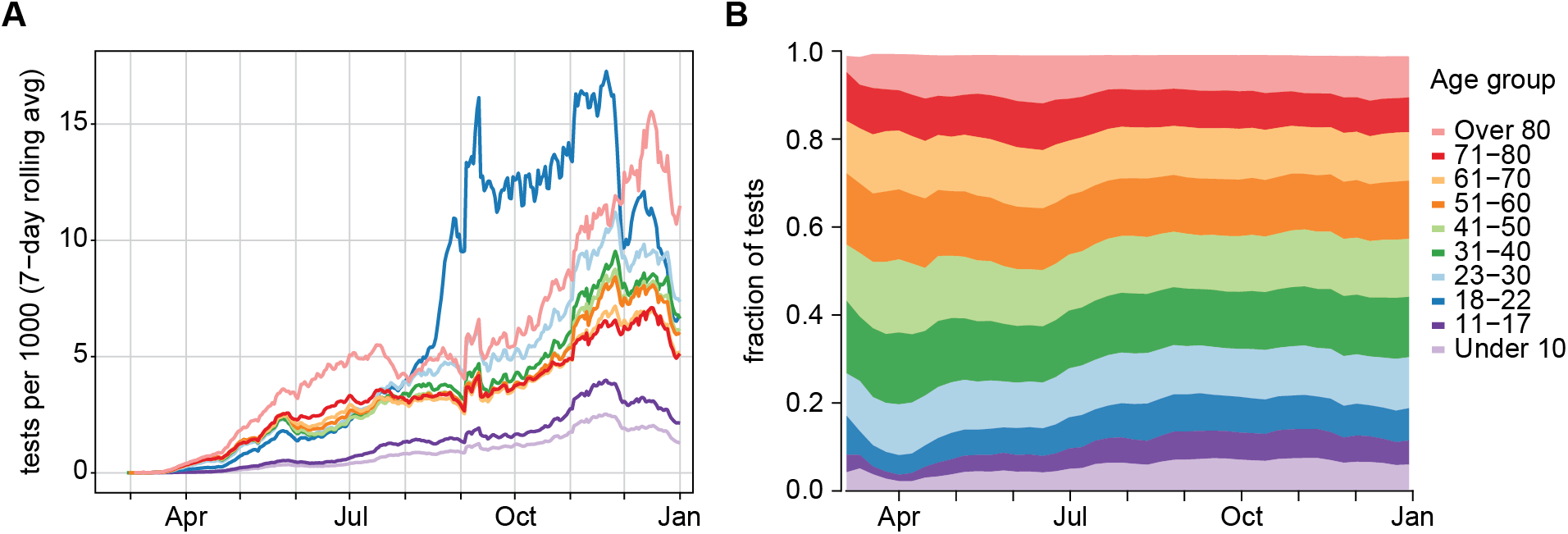
SARS-CoV-2 diagnosic testing in Illinois stratified by age group. (A) Seven-day moving average of daily tests per 1,000 population, by age group. (B) Share of tests by age group.

The portion of tests that were conducted on young people steadily increased between July and September 2020 (Figure 5B). Because COVID-19 is more likely to be asymptomatic or mild in younger patients (18,19), this change in the tested population would be expected to lead to lower case fatality rates in the population as a whole even without any improvements in treating COVID-19.

We compared the per capita testing rates of non-Hispanic White, Black, and Hispanic/Latino populations in Illinois (Figure 6). Testing increased between March and October for all three groups. During the first wave, per capita testing was highest in Black and Hispanic/Latino populations for most age groups, reflecting their disproportionate share of COVID-19 burden (14). After the end of the first wave in late June, testing was consistently lowest in Hispanic/Latino populations, with only minimal expansion of testing among Hispanic/Latino elders. The testing rate in the Black population saw a sharp increase around the beginning of July but did not increase further between July and October. In contrast, the testing rate in the White population increased steadily. The impact of student testing at university campuses was visible in all three demographic groups as the step-increase from mid-August to late-November and was largest in the White population. Testing rates began to decline sharply in all groups by mid-December.

**Figure 6.**
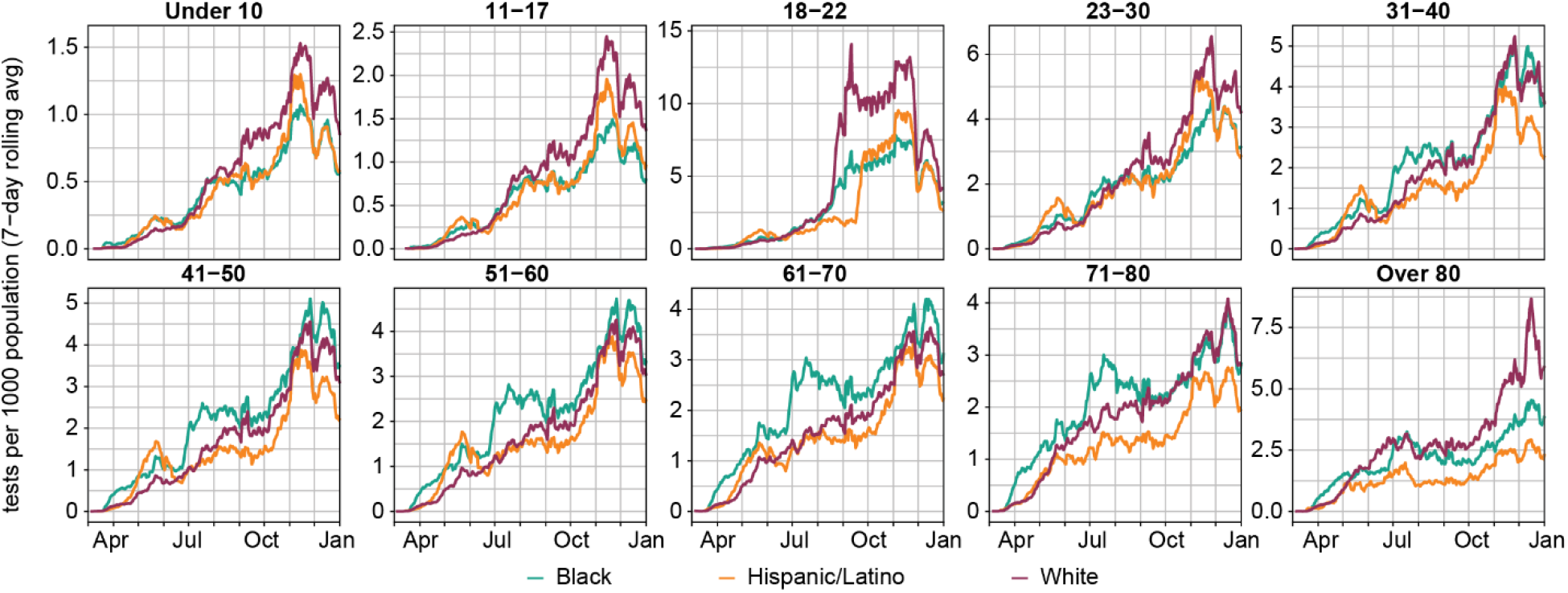
Seven-day moving average of daily SARS-CoV-2 tests per 1,000 population, by race/ethnicity and age group in Illinois in 2020.

On a per capita basis, testing intensity was lowest in Hispanic-Latino populations and highest in Black populations, although the extent of this difference appeared to vary by COVID-19 Region (Figure S2). Since the pandemic has disproportionately affected Black and Hispanic/Latino communities in Illinois (14), the higher testing intensity in Black populations does not necessarily mean that the testing was sufficient to capture burden relative to White populations.

### Crude assessment of under-testing with case fatality ratio

We considered whether the naive case fatality ratio (CFR), defined as the number of COVID-19 deaths divided by number of detected cases, could assess differences in SARS-CoV-2 testing in each super-region when detailed data on testing rates and hospital admissions were unavailable. Assuming that COVID-19 death ascertainment rates were uniformly high, and there was little to no geographic variation in infection fatality rate for SARS-CoV-2, we expected areas and populations with higher testing rates to also have lower case fatality rates.

In Illinois, crude CFR decreased over the course of 2020 in all super-regions and age groups (Figure 7), concurrent with the scale-up of testing. CFR was highest in the Northeast and Southern super-regions in the first two months of the epidemic, although CFR in the Central super-region increased in May and June for adults aged 41-50 years and 61-70 years. From July onward, CFR was similar in all regions.

**Figure 7.**
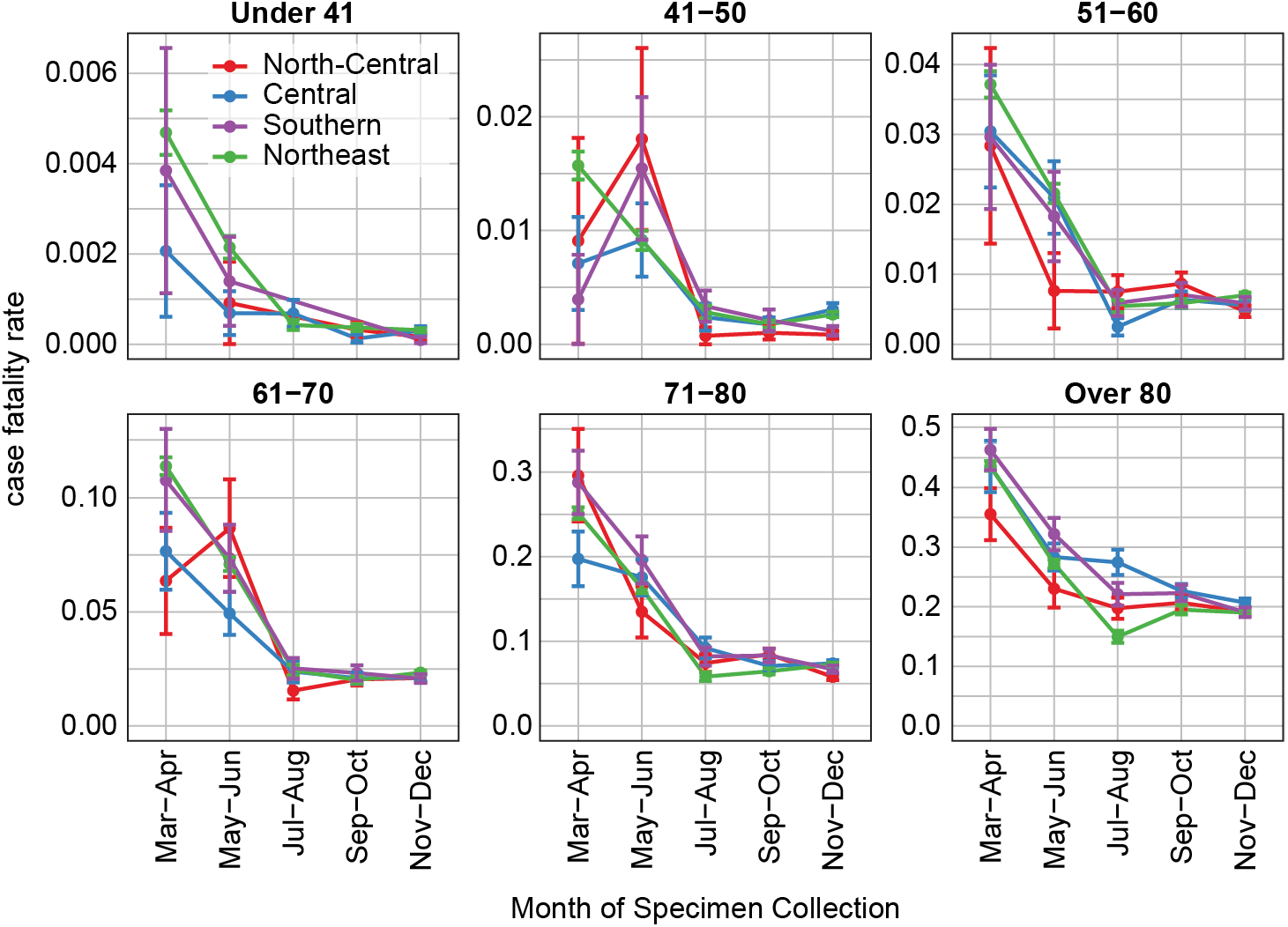
COVID-19 case fatality ratios (CFR), the fraction of recorded cases with a COVID-19-attributed death, by age group and super-region, for cases detected in 2020. Error bars indicate the standard error in regional CFR.

Differences in CFR could be driven by heterogeneous access to testing and care as well as regional differences in the prevalence of comorbidities, standard of care, or hospital capacity. The higher CFR in the Southern super-region prior to July reflected the consistently lower testing rates in COVID-19 Regions 4 and 5 during the first wave. However, the Northeast’s CFR prior to July was substantially elevated over the CFRs in the Central and North-Central super-regions despite the Northeast super-region’s higher intensity of testing. This discrepancy suggested that despite its higher testing intensity during the first wave, the Northeast super-region was disproportionately under-tested relative to its share of the state’s COVID-19 burden. Alternatively, the higher CFR despite higher testing in the Northeast could have been driven by insufficient targeting of tests to the most affected populations. The convergence of regional CFRs in late 2020 suggested that earlier differences in the regional CFRs might not have been driven by differences in regional prevalence of comorbidities. In April-May 2020, the Northeast super-region experienced the greatest strain on hospital capacity (Figure S3), which could have contributed to lower quality of care. However, hospital capacity overall is also highest in the Northeast, and peak inpatient census did not exceed capacity (Table S1).

Most majority-Black and majority-Hispanic/Latino ZIP codes are in the Northeast and Southern super-regions (10), where the highest overall CFRs were observed during March-April 2020. When stratified by age and race (Figure 8), CFR increased with age, with disparities becoming less pronounced for older age groups and as the epidemic progressed. CFRs remained higher for Black and Hispanic populations for all but the over-80 year-old age group through the end of 2020. Higher fatality rates among Black and Hispanic/Latino cases could be due to a combination of under-testing leading to fewer detected cases and disparities in clinical outcomes resulting in more deaths. The under-testing of Black and Hispanic/Latino populations is reflected in the higher CFRs in these populations. While elevated prevalence of comorbidities such as diabetes and hypertension increased the underlying infection fatality rate in in Black and Hispanic/Latino populations, the enormous difference in CFR in younger age groups in Mar-Jun 2020 is unlikely to be explained by comorbidities alone. For example, if diabetes prevalence at age 45 were around 5% in non-Hispanic Whites and 11% in non-Hispanic Blacks (20), and diabetes increased the risk of severe outcomes by around 60% (21), the disparity in diabetes prevalence would increase the CFR in the Black population ages 41-50 by approximately 10%. Yet in this age group, the relative risk of death given a case was almost 300% higher in the Black population compared to White in Mar-Apr 2020 [2.94 (95% CI: 1.72-5.03)]. In Nov-Dec 2020, relative risk remained high at 3.31 (2.25-4.89).

**Figure 8.**
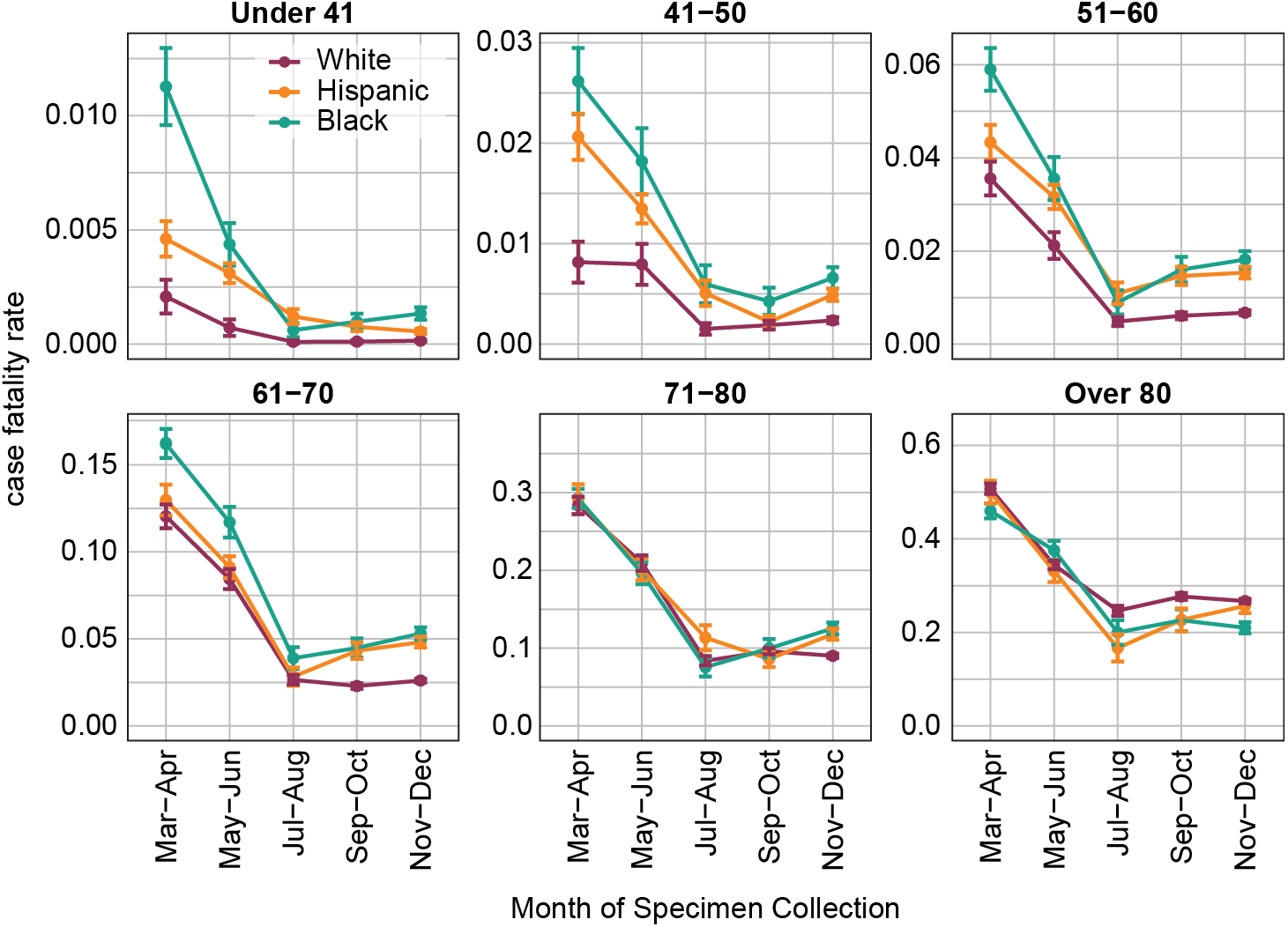
COVID-19 case fatality ratio by age group in non-Hispanic White, Hispanic/Latino, and non-Hispanic Black populations, for cases detected in 2020. Error bars indicate the standard error in group CFR.

In a sensitivity analysis, similar results were observed when cases with “unknown” ethnicity were removed altogether, instead of being allocated to non-Hispanic racial groups (Figure S4). As presented, the latter scenario may slightly underestimate CFR for older Black and White populations.

### The majority of SARS-CoV-2 infections were never detected

Low per capita testing rates are not necessarily problematic if there is little SARS-CoV-2 circulation and testing is highly targeted. These conditions do not describe Illinois in 2020. To estimate the extent to which testing was able to identify all incident SARS-CoV-2 infections in Illinois as a whole, we compared the expected IFR among individuals aged 61-70 years, as estimated by Levin et al. (11) and O’Driscoll et al. (12), to the CFR among the same group (Figure 9A). We restricted the analysis to this age group because detection rates are likely highly heterogeneous across age groups, and the 61-70 age group has a sizable number of weekly cases and deaths. We generated a naive estimate (Figure 9D) as well as estimates accounting for both a non-stationary IFR due to improving clinical outcomes among the infected (Figure 9B, E) and under-reporting of deaths (Figure 9C, F). This methodology only provides estimates of the detection rate for all infections and does not account for any heterogeneity in the detection of mild symptomatic and asymptomatic infections versus severely symptomatic infections. Because O’Driscoll et al. (12) estimates a lower IFR for ages 61-70 than Levin et al. (11), the estimated fraction of infections detected using IFR from O’Driscoll et al. is slightly lower than the same estimate using IFR from Levin et al.

**Figure 9.**
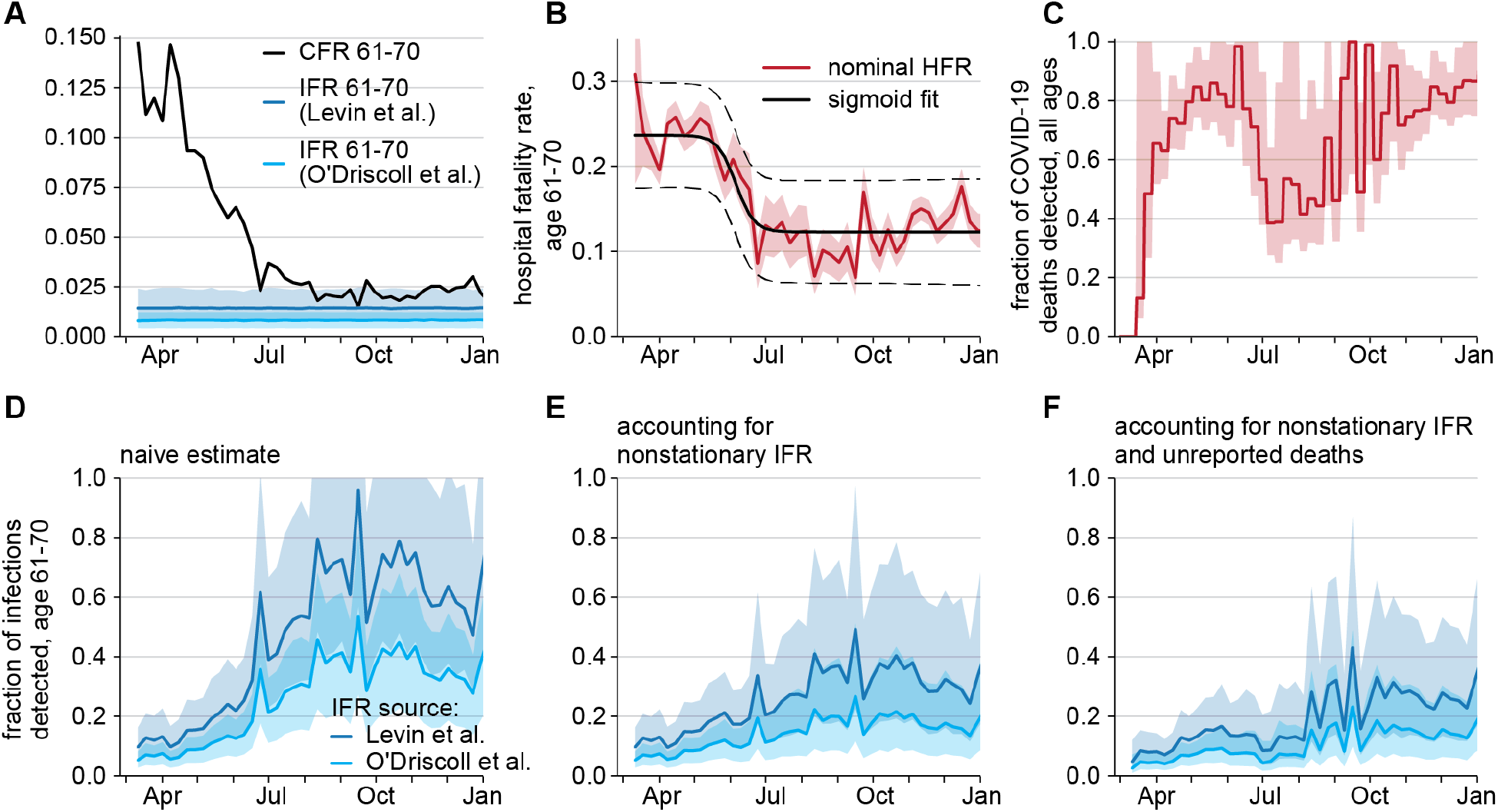
Estimating fraction of infections detected. (A) Naive CFR among adults aged 61-70 years by week of specimen collection alongside expected IFR among this age distribution, as estimated by Levin et al. (11) or O’Driscoll et al. (12). Shaded regions show 95% confidence intervals. (B) HFR among admitted 61-70 year-olds by week of specimen collection (solid red) with standard error of proportions (shaded red), fitted sigmoid curve (solid black) and 95% prediction interval (dashed black). (C) Fraction of all deaths reported by date of death, based upon comparison of COVID-19 mortality and excess all-cause mortality. (D) Estimated fraction of SARS-CoV-2 infections among adults aged 61-70-years that were detected by surveillance: assuming that all deaths are reported and IFR is stationary; (E) assuming all COVID-19 deaths are reported and IFR is non-stationary due to improving clinical outcomes; (F) and assuming that excess deaths are unreported COVID-19 deaths and IFR is non-stationary. Dark blue estimates use IFR estimates from Levin et al. (11) and light blue from O’Driscoll et al. (12). Shaded regions in D-F are 95% credible intervals with 1000 bootstrapped samples.

In March and April 2020, excess deaths in Illinois greatly exceeded COVID-19-attributed deaths, driving down the estimates of infection detection rates in Figure 9F. Due to lack of data on cause of death, we made the simplifying assumption in Figure 9C and 10F that all excess select-cause deaths documented by NCHS were COVID-19 related. This assumption produced a floor on the estimated detection rate of SARS-CoV-2 infections because not all excess deaths would be directly related to COVID-19. The true fraction of SARS-CoV-2 infections detected in Illinois likely lay between the estimates in Figure 9E and 9F.

In the early epidemic, prior to mid-April, we estimated that less than 10% of SARS-CoV-2 infections among adults aged 61-70 years were detected and reported to IDPH (Figure 9D-F). This low level of detection in the early stages of the epidemic is consistent with other estimates of around 10% detection rate (3,22). Despite the 3- to 4-fold scale-up in testing volume over the summer, we estimated that the detection rate among this population had yet to exceed 40% as of late December and could have been as low as under 20%.

Compared with the 61-70 year old age group (Figure 9), we expected the detection rate of SARS-CoV-2 infections in individuals over 70 years old to be higher (22). Older people are more likely to show symptoms and thus seek testing, testing intensity generally increases with age, and routine testing in long-term care facilities may additionally identify asymptomatic infections. Infections among younger age groups might have been detected at a lower rate than that of the 61-70 age group because younger adults were less likely to present symptoms. An exception was in college-age young adults: routine testing by universities could have led to higher overall detection rates than 40% in this population, although the overall rate would have masked heterogeneity across the state and in different segments of the college-age population.

## Discussion

Since the first cases of SARS-CoV-2 infection were detected in Illinois toward the end of January 2020, diagnostic testing capabilities have expanded dramatically. At the epidemic peak in November 2020, Illinois conducted over 110,000 diagnostic tests per day, among the highest in the U.S. For most of 2020, testing intensity varied considerably across the state, with the lowest rates in the Southern super-region and the highest rates in Champaign County, where UIUC rolled out mass testing in preparation for students returning to campus. Assessing trends in testing at the state level is insufficient as it masks local heterogeneities that can be critical: for example, UIUC’s testing protocols were not representative of testing protocols throughout the state, and any testing data, including cases, positive tests, and test positivity rate, from Champaign County or containing the college-age students would skew overall observed trends. While stratification of testing data by modality (inpatient, outpatient symptomatic, possible exposure, or routine) would permit disambiguation of apparent trends and more complete assessment of access to testing, these data were not systematically reported.

The ramp-up of testing, while necessary and impressive, is unlikely to be sufficient to contain SARS-CoV-2 on its own (6,23,24). Among individuals aged 61-70 years in Illinois, we estimated that as of mid-September, no more than 40% of all new infections were detected. Unfortunately, data on symptoms and reason for test were lacking. Because the majority of infections in the 61-70 year age group are likely to have been symptomatic (25,26), our estimated ceiling of 40% suggests that in addition to few asymptomatic cases found, there was also considerable room for improvement in the detection of symptomatic cases. Given RT-PCR sensitivity, some of the positive tests might have been old infections past their peak infectiousness period (27). Test turnaround times were often several days or more (28).

Modeling analyses have suggested that infections would need to be detected at a rate far greater than 40%, the high end of our Illinois estimates, for diagnostic testing to have had a substantial impact on containing transmission, even with all identified infections successfully isolated and test turnaround time within 2 days (23,29,30). Universal routine testing every 2 weeks with a highly sensitive diagnostic, a testing regime wherein nearly all infections would be identified, coupled with isolation of those who test positive, would reduce transmission by only 30% (23). Overall, diagnostic testing probably played a minor role in directly reducing SARS-CoV-2 spread in Illinois, second to other mitigation measures (e.g. social distancing, mask usage, retail/restaurant closures). Because Illinois testing rates have been among the highest in the United States (31), diagnostic testing likely had minimal direct impact on reducing SARS-CoV-2 spread in most U.S. states in 2020. However, diagnostic testing also has an indirect role in reducing transmission by providing surveillance data, allowing the public to take preventive measures and policymakers to make mitigation policy decisions.

The ramp-up of testing did not result in equity of access to diagnostic testing sites, as many residents of Illinois outside the Northeast metro area, particularly residents of Central and Southern Illinois, needed to travel many miles to the nearest testing site. The disparities between urban, suburban, and rural access to testing were likely to be similar in other parts of the U.S.

Current surveillance does not give us an accurate picture of spread in different populations within a state. Race and ethnicity data were missing for over 40% of tests. More testing and more complete demographic and epidemiologic data are needed to capture cases all over Illinois, and particularly in racial and ethnic minority populations who experience higher rates of occupational exposure and reduced access to care (32,33). Black populations were under-tested for SARS-CoV-2 in Utah (34) and New York City (35). Black patients were more likely to access testing in hospitals rather than outpatient settings in California (36), which could have reflected limited access to ambulatory testing sites or decisions to not undergo testing unless or until symptoms became severe. In a cohort study of people receiving care through the U.S. Department of Veterans Affairs, Black and Hispanic patients had both higher rates of testing and higher rates of positivity (37). Disparity in surveillance quality across socioeconomic strata, where communities that experience disproportionate risk also have the poorest quality surveillance, is hardly unique to COVID-19 (38).

If detailed testing data are unavailable, we found that CFR can act as a crude benchmark of relative under-testing across geographic regions and reflects disparities in testing across demographic groups. However, CFR-based indicators of under-testing should be used with caution, as multiple mechanisms can create differences in observed CFR across populations, and cumulative CFRs may not reflect current conditions.

Illinois exerted tremendous effort to scale up diagnostic testing and successfully reduce geographic and demographic disparities in testing rates. Yet, containment through diagnostic testing alone would have required another order of magnitude increase in testing capacity. Managing the COVID-19 pandemic in the U.S. requires an integrated strategy of multiple policies and interventions, of which testing is only one part.

Testing is critical both as an intervention, as positive cases are directed to isolate and prevent transmission, and for surveillance, which provides information for policymakers to make effective decisions. However, when testing is both insufficient and heterogeneous, existing inequalities in disease burden are exacerbated, surveillance quality suffers, and directing interventions to appropriate demographics and locales becomes challenging. Understanding the disparities in testing is the first step toward building surveillance structures capable of reliably informing good decisions. Identifying which geographic areas are relatively under-tested with the straightforward methods as demonstrated here can be a critical part of public health departments’ regular assessment of their testing capacity.

## Conclusions

In the U.S. state of Illinois, testing intensity continues to vary geographically and across demographic groups. While testing rates improved dramatically from the onset of the pandemic through December 2020, the Southern and Central parts of the state remained relatively under-tested. These data suggest that raw per capita testing volume, infection detection rates derived from deaths and IFR, CFR, and disparate patterns in admissions and cases can all be used to identify populations in which testing should be expanded. By accessing a variety of available data sources, policymakers can strengthen their understanding of COVID-19 disease burden throughout the state to more accurately assess where to target additional testing resources, thus strengthening the potential for infected individuals to be safely isolated and referred to appropriate care.

## Data Availability

The I-NEDSS and testing by age and race/ethnicity datasets analyzed in this study were used under license for the current study, and so are not publicly available. Restrictions apply to the availability of these data, which contain identifiable private health information. Interested parties should contact IDPH to inquire about access. Public data on cases and testing are available from IDPH (https://www.dph.illinois.gov/covid19/covid19-statistics) and from other public aggregators (https://coronavirus.jhu.edu/region/us/illinois).

https://coronavirus.jhu.edu/region/us/illinois

## List of abbreviations

ACS: American Community Survey
CFR: case fatality ratio
HFR: hospital fatality ratio
IDPH: Illinois Department of Public Health
IFR: infection fatality ratio
I-NEDSS: Illinois’s National Electronic Disease Surveillance System
NCHS: National Center for Health Statistics
UIUC: University of Illinois at Urbana-Champaign

## Declarations

### Ethics approval and consent to participate

This study was carried out as part of a Medical Study “Modeling COVID-19 Epidemiologic Trend and Health Care Impact in Illinois” declared by IDPH on March 23, 2020. All data collection was performed by IDPH as part of routine surveillance for COVID-19 and was deidentified prior to analysis.

This activity was reviewed by CDC and was conducted consistent with applicable federal law and CDC policy. See e.g., 45 C.F.R. part 46, 21 C.F.R. part 56; 42 U.S.C. §241(d); 5 U.S.C. §552a; 44 U.S.C. §3501 et seq.

### Consent for publication

Not applicable

### Authors’ contributions

JG and SC conceived the project. TMH completed the race and ethnicity analysis. RAKR completed the testing site and infection detection rate analysis. PA contributed to the detection rate analysis. TMH and EW completed the CFR analysis. JG and MR completed the regional testing rate analysis. MR completed the bed availability analysis. TMH, RAKR, and JG wrote the initial draft. All authors revised and approved the final manuscript.

## Acknowledgements

We thank Stacey Hoferka Jensen, Dejan Jovanov, and Sara Rogers for data extraction and preparation from I-NEDSS, and Arielle Eagan for comments on the manuscript.

## Disclaimer

The findings and conclusions in this report are those of the authors and do not necessarily represent the official position of the Centers for Disease Control and Prevention.

---

**Figure S1.**
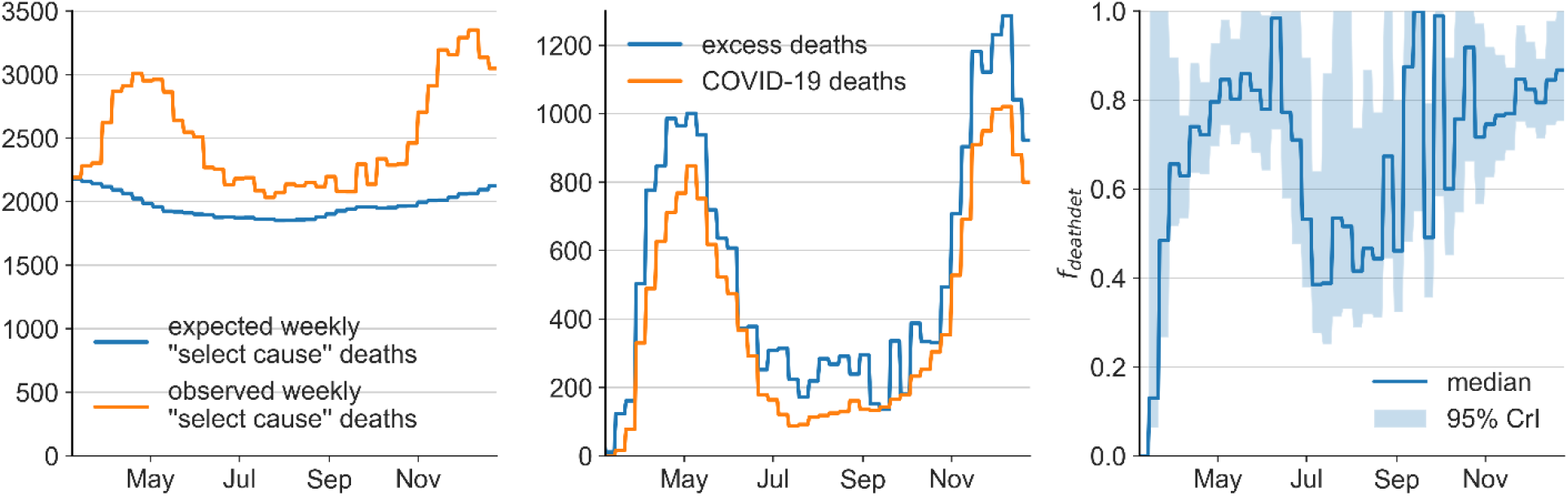
Calculation of *f*_*deathdet*_ from Illinois COVID-19-attributed deaths and excess select cause deaths.

**Figure S2.**
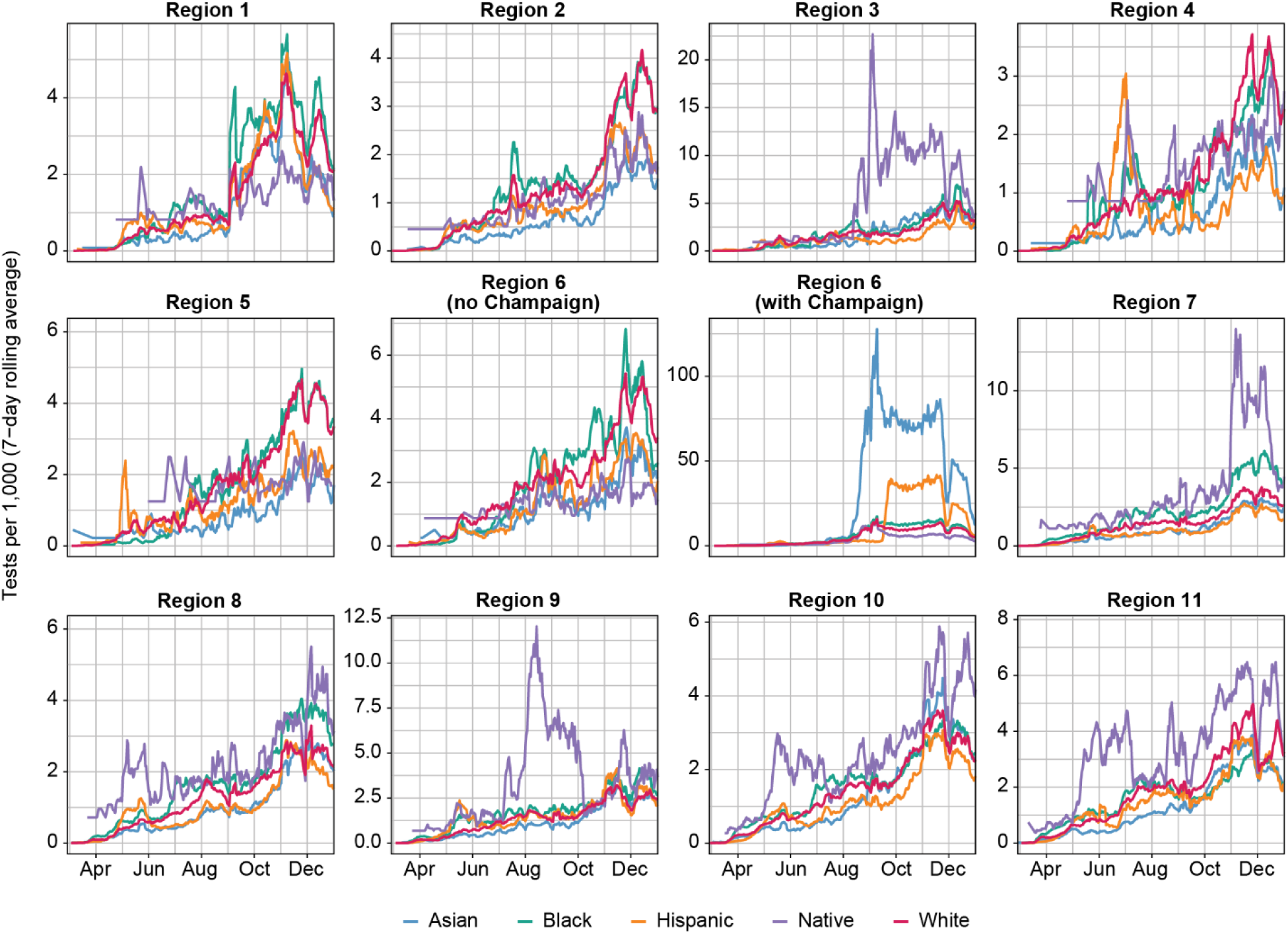
Testing rate per capita by race/ethnicity and COVID-19 Region. Population denominators were drawn from ACS 2018. “Native” included both “American Indian or Alaskan Native” and “Native Hawaiian or Other Pacific Islander”. Asian and Native groups were excluded from Figures 6 and 8 due to small population denominators after stratifying by age. The small denominators are reflected in the large fluctuations in the Native timeseries in this figure.

**Figure S3.**
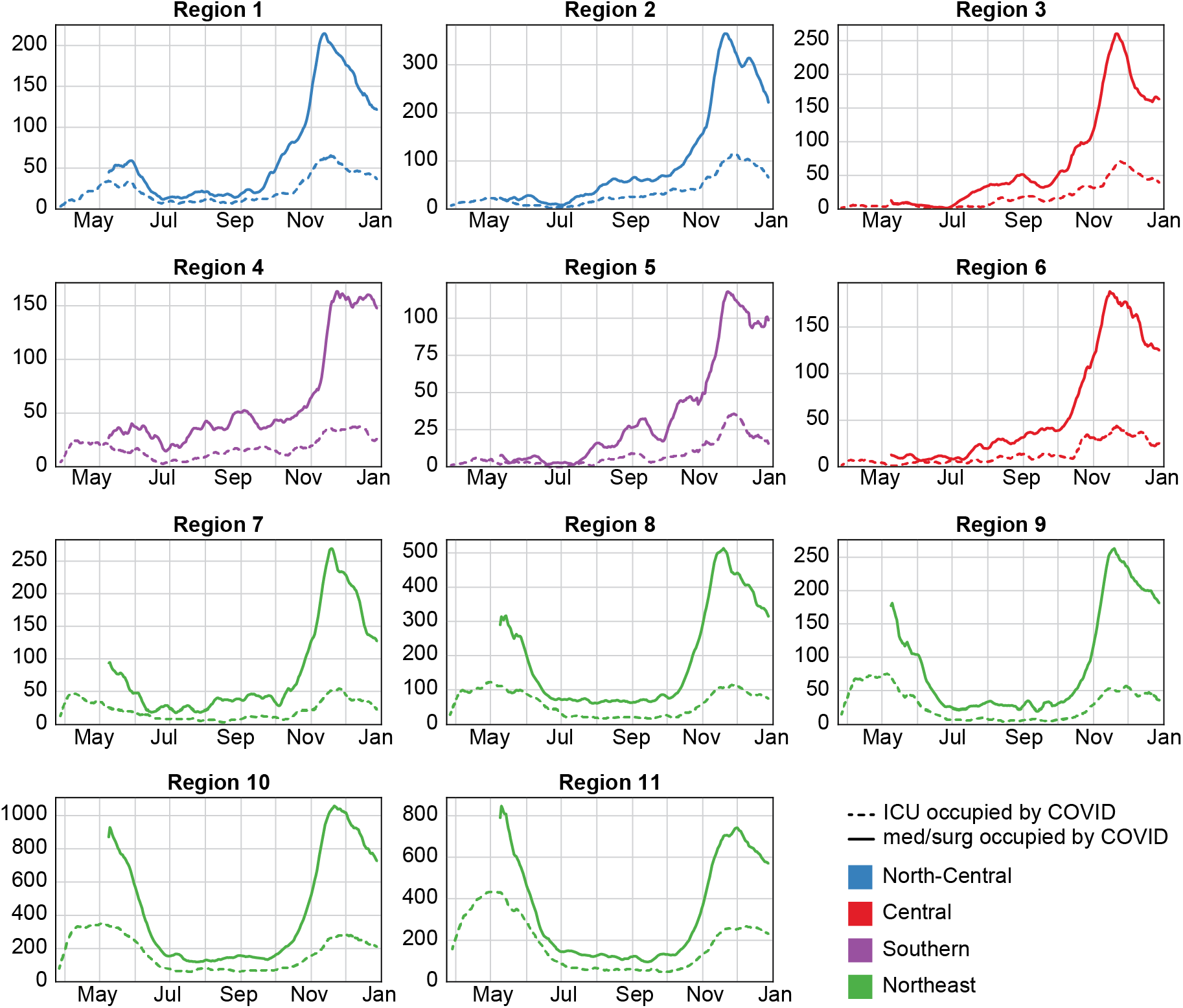
Daily COVID-19 hospital census by COVID-19 Region (subplots) and super-region (colors) in 2020. Lines show 7-day rolling averages. Data for med/surg occupancy are not available prior to May 2020. Med/surg census covers all admitted patients who are not in the intensive care unit (ICU).

**Figure S4.**
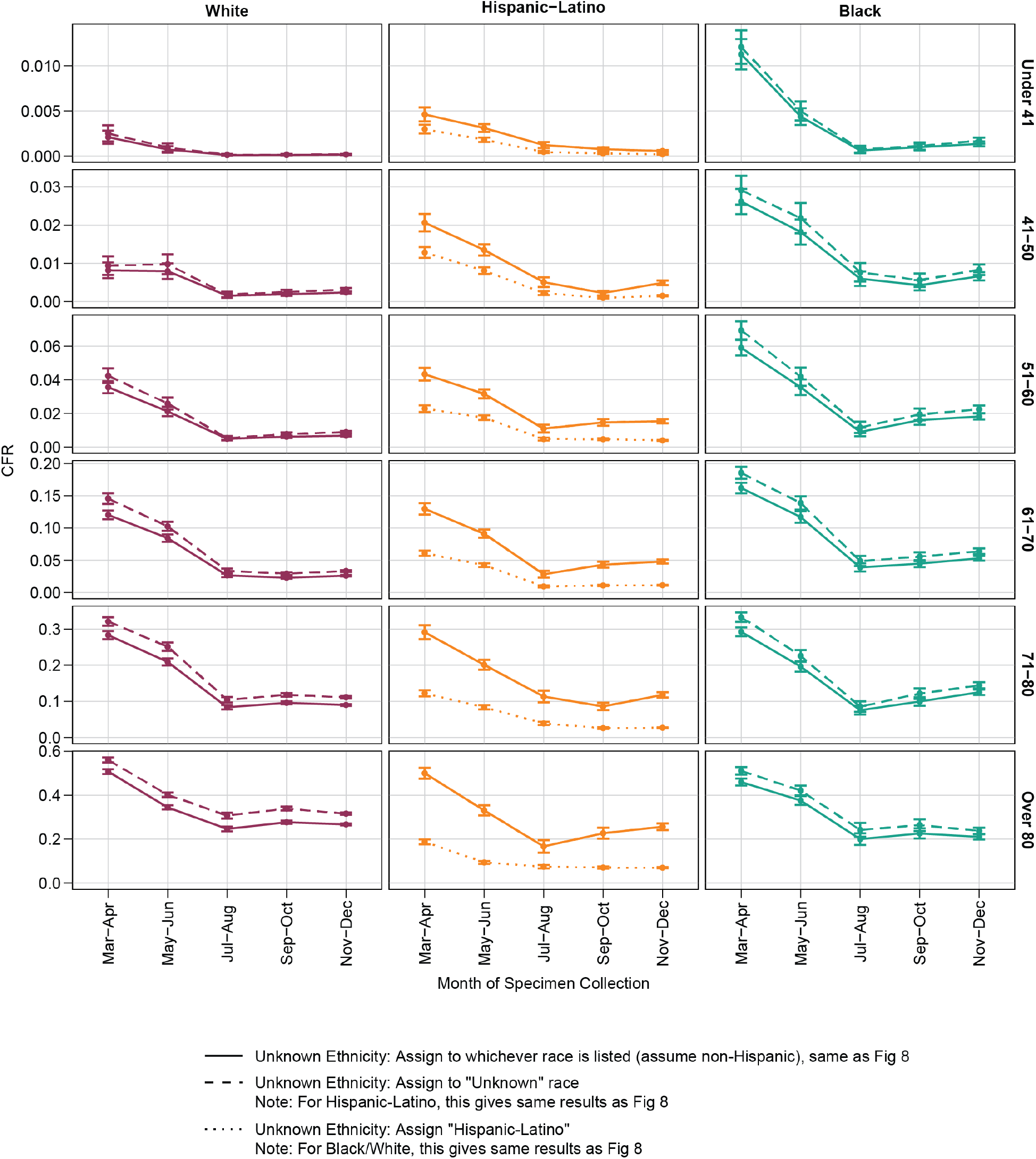
Sensitivity of CFR to case race/ethnicity assignment. Alternate scenarios were run in which cases with “unknown” ethnicity were: 1) Assumed to be non-Hispanic and assigned to the recorded racial group (solid line, and identical to Figure 8), 2) Assigned to the “unknown” racial group (dashed line), or 3) Assumed to be Hispanic (dotted line). For Hispanic-Latino, scenarios #1 and #2 are identical. For Black and White, scenarios #2 and #3 are identical. Assumption of non-Hispanic ethnicity slightly, but significantly, decreases CFR for older non-Hispanic White and Black populations.

**Table S1.** Total hospital beds by COVID-19 Region. Bed availability data from September 1, 2020 via IDPH website. Beds available for COVID refers to number of beds not occupied by non-COVID patients.

| COVID-19 Region | population | Med/Surg total beds | ICU total beds | Med/Surg beds available for COVID | ICU beds available for COVID | Med/Surg beds available for COVID per 10,000 | ICU beds available for COVID per 10,000 |
| --- | --- | --- | --- | --- | --- | --- | --- |
| 1 | 660965 | 939 | 229 | 428 | 124 | 6.48 | 1.88 |
| 2 | 1243906 | 1876 | 323 | 870 | 156 | 6.99 | 1.25 |
| 3 | 556776 | 1141 | 150 | 475 | 73 | 8.53 | 1.31 |
| 4 | 656946 | 862 | 130 | 343 | 86 | 5.22 | 1.31 |
| 5 | 403659 | 668 | 99 | 370 | 65 | 9.17 | 1.61 |
| 6 | 739098 | 1087 | 178 | 461 | 112 | 6.24 | 1.52 |
| 7 | 800605 | 820 | 174 | 320 | 63 | 4.00 | 0.79 |
| 8 | 1455324 | 1906 | 431 | 761 | 222 | 5.23 | 1.53 |
| 9 | 1004309 | 1117 | 254 | 513 | 140 | 5.11 | 1.39 |
| 10 | 2693959 | 3680 | 772 | 1237 | 354 | 4.59 | 1.31 |
| 11 | 2456274 | 4475 | 1066 | 1526 | 516 | 6.21 | 2.10 |

